# Prevalence and characteristics of women with polycystic ovary syndrome and isolated diagnostic feature using the International evidence-based guideline and antimüllerian hormone: A nationwide cross-sectional survey

**DOI:** 10.1101/2025.07.31.25332503

**Authors:** Md Shahed Morshed, Mashfiqul Hasan, Tania Tofail, Hurjahan Banu, Nusrat Sultana, Samirah Binte Illiyas, Munira Afroz Siddika, Rafayel Islam, Hafsa Mahrukh, Tahmina Ferdousi, Indira Roy, Yasmin Aktar, Hilary Yasmin, Farhana Rahman, Nazia Akter, Abdullah Al Noman Bhuiyan, Krishna Airi, Rifat Hossain Ratul, Prodipta Chowdhury, Koushik Ashraf, Rayhan Ali Mollah, Muhammad Abul Hasanat

**Affiliations:** PCOS study group, Department of Endocrinology, Bangladesh Medical University, Dhaka, Bangladesh

**Keywords:** Polycystic ovary syndrome, Antimullerian hormone, Prevalence, Irregular menstrual cycles, Hirsutism

## Abstract

**Background:** Previous studies reported polycystic ovary syndrome (PCOS) prevalence using consensus guidelines and ultrasonography. Data on stepwise decision-making by the International evidence-based guidelines and using anti-müllerian hormone (AMH) for prevalence were limited.

**Objective:** This nationwide cross-sectional survey determined the prevalence and characteristics of PCOS and isolated diagnostic features among reproductive-aged Bangladeshi women (10-45 years) using the International Evidence-based Guidelines, 2023, incorporating AMH.

**Methods:** Conducted April-September 2024 across eight divisions (one urban & one rural) of Bangladesh, 1201 females were sampled. Participants underwent fasting, OGTT, and detailed data collection (demographic, lifestyle, reproductive history, and physical measurements). Blood was analyzed for glucose, lipid, and ALT. Total testosterone (TT) was measured for isolated irregular cycles. Adults with only significant hirsutism or irregular cycles with normal testosterone levels were further evaluated by AMH. Thyroid dysfunction and hyperprolactinemia were excluded. TT and AMH cut-offs were derived from 100 hirsutism-free adult women with regular cycles.

**Results:** Of 1201 participants, 403 were excluded. Among 798 eligible women, 38 (4.8%) had both irregular cycles and significant hirsutism, 75 (9.4%) had only significant hirsutism, 141 (17.7%) had only irregular cycles, and 544 (68.2%) had neither. After TT and AMH evaluation and excluding two hyperprolactinemia cases, a 6.9% (55/798) prevalence was found among 57 women with probable PCOS. Familial predisposition, unhealthy sleep behaviour, and higher androgenic and metabolic features were observed in women with PCOS versus controls. Metabolic syndrome frequency was higher among adult PCOS (33.3% vs. 4.0%) than adolescent PCOS.

**Conclusion:** The community-based prevalence of PCOS among Bangladeshi women was 6.9%, with distinctive features compared to controls. Women with isolated diagnostic criteria require further evaluation and long-term follow-up.

**Highlights:** This study used a stepwise decision-making approach and antimüllerian hormone to determine the prevalence of polycystic ovary syndrome and isolated diagnostic features in a nationwide cross-sectional survey.

The estimated prevalence of PCOS was 6.9% among 798 reproductive-aged Bangladeshi women, consisting of 5.9% and 8.0% of adolescent and adult PCOS, respectively.

This study also identified 15.5% and 9.1% of women with isolated irregular cycles and significant hirsutism, respectively, who required further evaluation and follow-up.

## Introduction

Polycystic ovary syndrome (PCOS) is a prevalent yet complex endocrine and metabolic disorder affecting women of reproductive age. It encompasses a variety of issues, including reproductive, dermatological, cardiovascular, metabolic, and psychiatric symptoms (WHO, 2025). However, the nonspecific nature of many features complicates the diagnosis. Due to its unclear pathogenesis, PCOS diagnosis has historically relied on consensus criteria, leading to a broad spectrum of reported prevalence rates (Joham AE, 2022). Of the three widely recognized criteria, the Rotterdam criteria—requiring two out of three components: ovulatory dysfunction, clinical and/or biochemical hyperandrogenism, and polycystic ovarian morphology (PCOM)—are the most accepted and have been endorsed by the International Evidence-based Guidelines since 2018 (Teede HJ, et al., 2018). These updated guidelines reflect an ongoing effort within the medical community to enhance the accuracy and inclusivity of PCOS diagnosis, incorporating new evidence and biomarkers to define the syndrome better. This guideline clarifies each criterion based on evidence and suggests a stepwise decision-making approach, prioritizing clinical features over diagnostic investigations. The definitions of irregular cycles are now based on gynecological age, and among the clinical features of hyperandrogenism, only hirsutism has been accepted (Teede HJ, et al., 2023). The requirement for investigations to diagnose PCOS has been reduced in clinical settings (Pace L, et al., 2024 & 2025). The application of these recommendations in a community setting has not yet been adequately investigated. Additionally, the latest 2023 guidelines have incorporated antimüllerian hormone (AMH) as an alternative to ultrasonography (USG) criteria for PCOS, due to several limitations of USG, particularly operator dependence and the evolving follicle number per ovary criteria with the advancement of the probe (Di Michele S, et al., 2025). However, the PCOM criterion is not currently recommended for adolescents (10–19 years) (Teede HJ, et al., 2023). Previous prevalence studies reported their findings based on the different consensus-based guidelines and using USG criteria. However, the prevalence of PCOS, as per the International Evidence-based Guidelines, 2023, especially when applying AMH, was not adequately documented.

Being one of the most densely populated nations with a predominance of metabolic phenotypes, the burden and characteristics of Bangladeshi women with PCOS require further exploration, especially when the population-based prevalence is largely unknown. This study aimed to determine the prevalence and characteristics of PCOS among Bangladeshi reproductive-aged women by utilizing the International Evidence-based Guidelines, 2023, which included measuring AMH instead of USG (in adult women only), through a nationwide survey. In addition, we also described two groups of women who presented with an isolated feature of PCOS.

## Methods

### Study design and participants

This nationwide cross-sectional survey was done in all eight administrative divisions of Bangladesh between April and September 2024. Two sites, one urban ward and one rural village, comprising a total of 16 sites, were selected using a multistage cluster sampling method. Then, from the list (Bangladesh Population and Housing Census, 2022) provided by each local government authority, 100 females aged 10–45 years residing at the specific site for at least one year were selected using systematic random sampling and invited to participate in the study.

### Ethical consideration

Informed consent or assent was obtained from each participant before enrollment in the study. The study protocol was approved by the Institutional Review Board (IRB) of Bangladesh Medical University (No. BSMMU/2024/3963, date: 02.04.2024).

### Study procedure

All participants were requested to come in an overnight fast (8–14 hours) on a specified date, at a designated location, and at a prefixed time. After taking a fasting blood sample, a 75-g oral glucose tolerance test (OGTT) was performed. During the 2-hour OGTT waiting period, participants’ demographic, socioeconomic, lifestyle, and reproductive histories were recorded, along with related physical measurements [height, weight, waist circumference (WC), blood pressure (BP), thyromegaly, hirsutism, acne, and acanthosis nigricans] by a team of postgraduate students and fellows of our Department.

Participants’ food habits, leisure time, and sleep patterns were assessed using a questionnaire from the STEPS survey (Riaz BK, et al., 2020). The physical activities were classified into low, moderate, and high intensity, with cut-offs based on the metabolic equivalent task (MET) of 600 and 3000 minutes per week (Mumu SJ, 2017). We defined irregular cycles according to the International Evidence-based Guidelines (2023). We used a modified Ferriman-Gallway (mFG) score of 4 or greater as a cut-off point for significant hirsutism (Teede HJ, et al., 2023). Body mass index (BMI) was calculated from height and weight, and classified separately for adolescents [CDC BMI percentile: <85^th^ percentile (lean), ≥85^th^ (overweight-obesity)] and adults [BMI (kg/m^2^): <23 (Lean), ≥23.0 (overweight-obesity)] (WHO, 2020). Central obesity was defined as a WC ≥ 70^th^ percentile for age and sex in adolescents and ≥80 cm in adults (WHO, 2020; Khadilkar A, 2014). For individuals under 13 years of age, the BP percentile was calculated using an online calculator that takes into account age, sex, and height. Metabolic syndrome-BP was defined as BP ≥90^th^ percentile for 10–12 years and ≥130 mmHg and/or> 85 mmHg for the rest of the participants (Codazzi V, 2024).

### Investigations

Glucose values were immediately measured at the collection sites using a semi-automatic biochemical analyzer (BAS-150 TS Plus, Labomed, Inc., Los Angeles, U.S.A) with the glucose-oxidase method. The separated serum from the fasting blood was transported in ice boxes to the BMU for preservation in a −70 °C freezer. Serum AMH, TSH, FT4, total testosterone (TT), and prolactin levels were analyzed in the Department of Microbiology and Immunology, BMU. Serum AMH was analyzed by Maglumi® AMH kit using chemiluminescence immunoassay (CLIA) with a detection limit of 0.02 – 25 ng/mL, intra-assay coefficient of variations (CVs) of 3.06% – 4.04%, and inter-assay CV of 0.83% – 2.56%. TT was measured using a similar kit and method, with detection limits, intra-assay CVs, and inter-assay CVs ranging from 0.25 to 17.0 ng/dL, 2.98% to 3.88%, and 4.43% to 6.27%, respectively. Other hormones were also analyzed by CLIA. Additionally, serum lipid profiles and alanine aminotransferase (ALT) levels were measured from fasting samples using an automated analyzer (Architect Plus ci8200) in the Department of Biochemistry and Molecular Biology of BMU.

As we have no population-specific data, we randomly chose 100 adult women (>19 years) with regular cycles and a mFG score of ‘0’ from our healthy participants to measure TT and AMH. The 95^th^ percentile values of TT (>53.2 ng/dL) and AMH (>4.2 ng/mL) were used as cut-offs for hyperandrogenemia and PCOM, respectively. Thyroid dysfunctions were defined by TSH levels below 0.1 or above 10 mIU/mL, and hyperprolactinemia was defined by prolactin levels above 52.9 ng/mL. The following abnormalities were defined as components of metabolic syndrome: waist circumference ≥80 cm or age-specific cut-offs, systolic BP ≥130 mm-Hg &/or diastolic BP ≥85 mm-Hg, fasting plasma glucose, FPG ≥5.6 mmol/L, HDL-cholesterol <50 mg/dL for adults and <40 mg/dL for adolescents, and triglycerides (TG) ≥150 mg/dL for all. The International Harmonization criteria were used to define metabolic syndrome (Codazzi V, 2024; Alberti, 2009).

### The stepwise decision making

Out of 1600 invited women, 1201 participated. Among them, 403 were excluded for various reasons. The remaining 798 women were evaluated for irregular cycles and significant hirsutism. Those who presented with only irregular cycles were initially analyzed for their TT levels. If it was normal, we then further evaluated them by AMH only in adult women. Similarly, in adults with only significant hirsutism, we measured their AMH levels. Those meeting two criteria — irregular cycle + significant hirsutism/hyperandrogenemia, irregular cycle + increased AMH (adults), and significant hirsutism + increased AMH (adults) — were further evaluated for TSH, FT4, and prolactin to exclude thyroid dysfunctions and hyperprolactinemia. The fulfilment of two criteria, along with the exclusion of similar endocrinopathies, makes a diagnosis of PCOS.

### Statistical analysis

Data were entered, edited, and analyzed using the SPSS software version 25.0. The participants’ socioeconomic status was divided into lower, middle, and upper based on the wealth index (WI). It was calculated using principal component analysis based on the household asset data. Qualitative data were expressed in frequency and percentages. The quantitative data were tested for distribution using the Shapiro-Wilk test. Quantitative data were expressed as the mean with standard deviation (SD) for normally distributed data and as the median with interquartile range (IQR) for skewed data. The comparison between two qualitative variables was analyzed using the Chi-squared test or Fisher’s exact test. In cases of significant association, cells with adjusted standardized residuals (ASRs) outside the range of ±3 were considered significant. The quantitative variables were evaluated with qualitative variables using the Mann-Whitney U test or the Kruskal-Wallis test (with post hoc pairwise comparison). Any two-tailed p-value below 0.05 was considered statistically significant.

## Results

### Prevalence of PCOS

We started with 1201 reproductive-aged women (10–45 years). After excluding 403 women for various reasons, 798 women were eligible for further evaluation. Among them, 544 (68.2%) had regular cycles and insignificant hirsutism (controls). According to the International Evidence-based Guideline, 179 had at least one type of irregular cycle. Isolated irregular cycles (IIC) were present in 141 (17.7%), isolated significant hirsutism (ISH) was present in 75 (9.4%), and both diagnostic features (probable PCOS) were present in 38 (4.8%). Those with only IIC were evaluated by TT. Hyperandrogenemia (TT > 53.2 ng/dL) was present in 12 patients. The adults with normoandrogenemia (n= 83) were then assessed by measuring AMH. PCOM, defined by AMH (>4.2 ng/mL), was present in five participants. Of 141 women with IIC, a total of 17 participants were labelled as probable PCOS. Among adult participants with ISH (n=29), only two had PCOM and were labelled as probable PCOS. Out of 57 possible cases of PCOS, two were excluded due to hyperprolactinemia (>52.9 ng/mL), and none had thyroid dysfunctions. Overall, 55 out of 798 [6.9% (95% CI: 5.2% to 8.9%)] had PCOS, including 25 (5.9%) adolescents and 30 adults (8.0%) (Figure 1).

**Figure 1:**
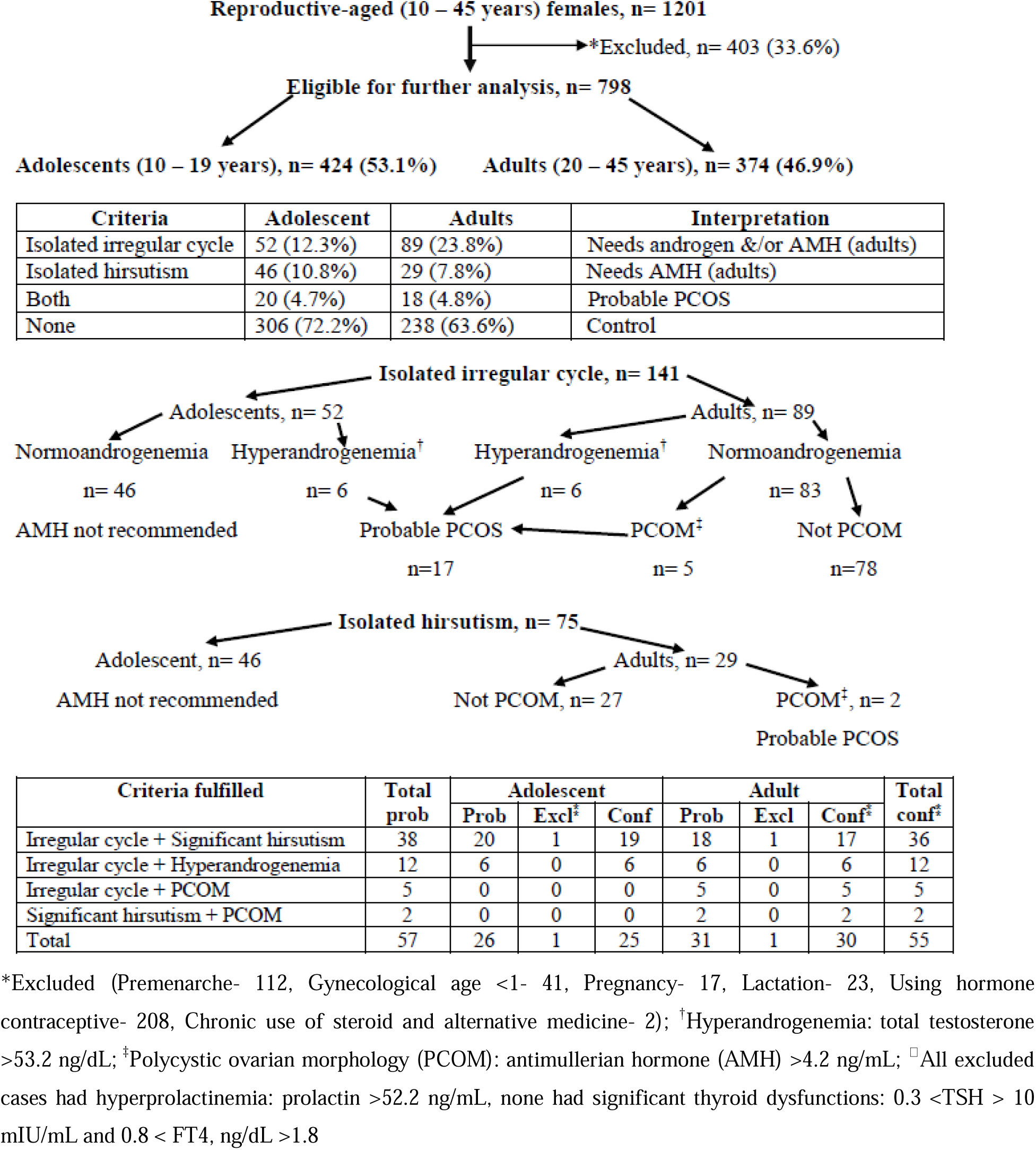
The stepwise diagnostic flowchart of PCOS

### Characteristics of the study groups

The characteristics of women with PCOS in comparison to controls, isolated irregular cycles, and isolated significant hirsutism are shown in Tables 1, 2, and 3. The analysis is limited to 796 participants, excluding the two cases of hyperprolactinemia. Those with IIC had a higher age than those with ISH (p=0.002) and control (p=0.001). There were significant differences in age groups (χ^2^=19.1, Cramer’s V, φ_c_= 0.2), residence (χ^2^=12.0, φ_c_= 0.1), division (χ^2^=65.6, φ_c_= 0.2), and occupation (χ^2^=20.9, φ_c_=0.1) among the study groups. In those with IIC, the frequency of adults was higher (ASR = +3.9). Among women with IIC, the frequency of housewives was higher (ASR=+3.6), but students were lower (ASR=-3.9). The prevalence of PCOS was higher in the Rajshahi (12.5%) and Dhaka (9.6%) divisions and lowest in the Chattagram (2.7%) and Khulna (2.2%) divisions. The frequency of participants from the Rangpur division was higher in the ISH (ASR = +4.0), but lower in the control group (ASR = - 3.7). Participants from the Mymensingh division were higher in the control group (ASR = +3.5) (Table 1).

**Table 1:**
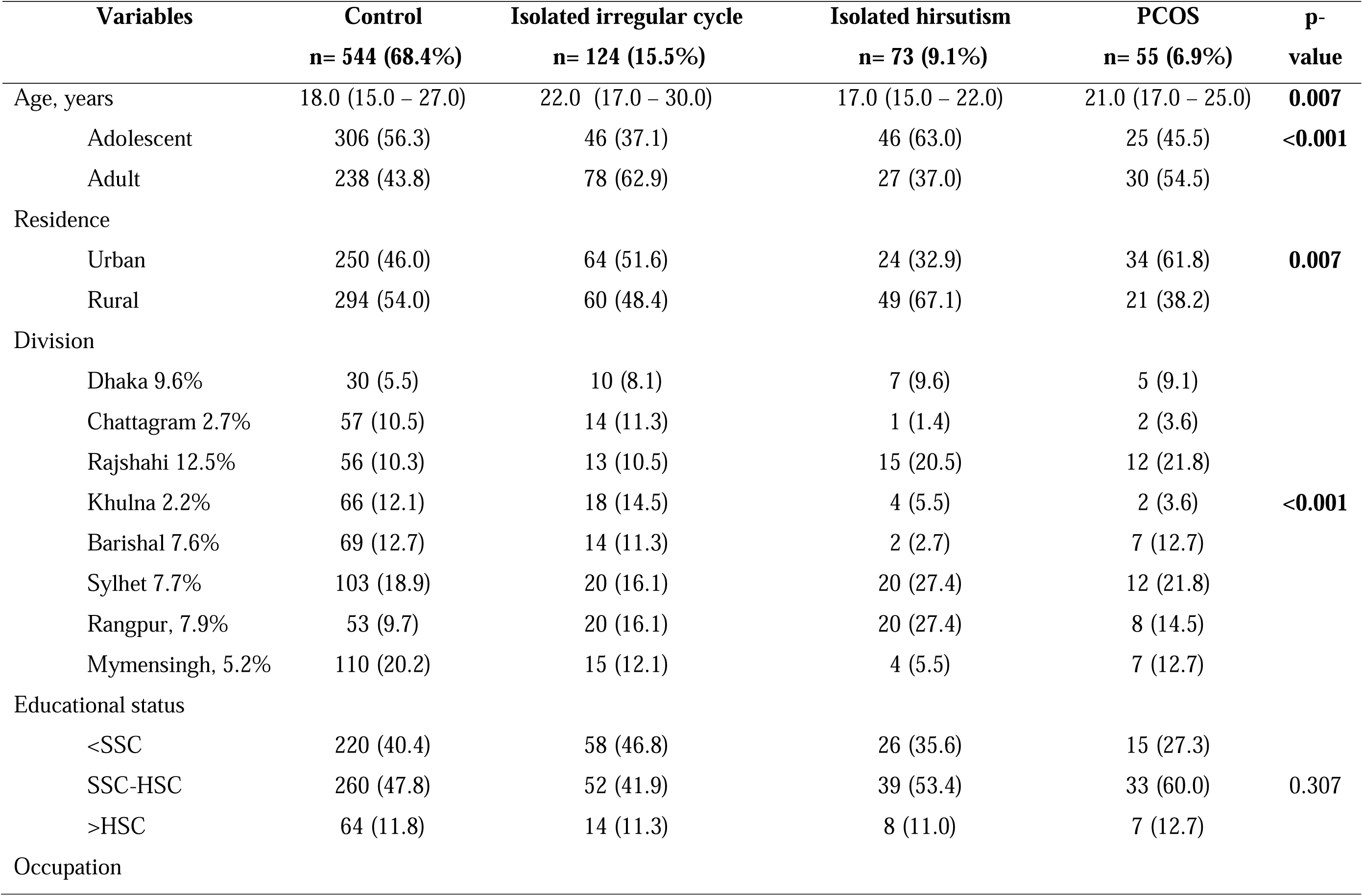
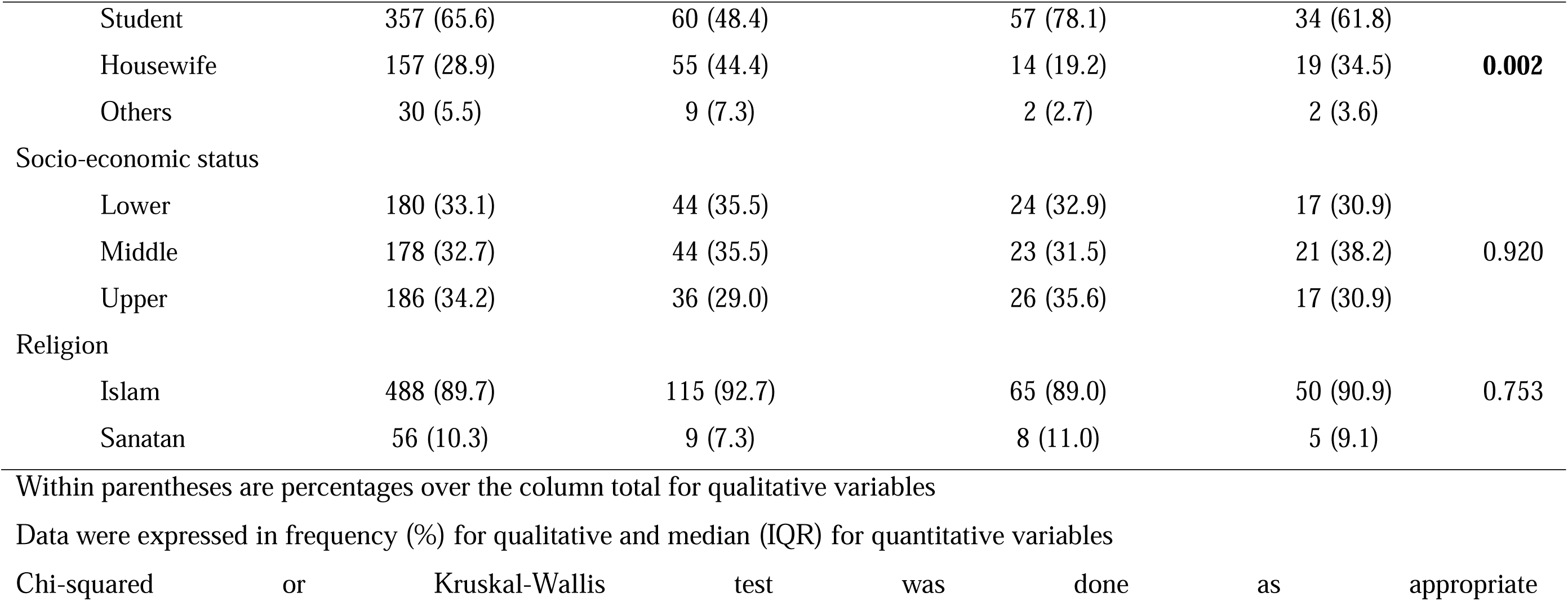
Socio-demographic characteristics of the study participants, n= 796.

| Variables | Control<br>n= 544 (68.4%) | Isolated irregular cycle<br>n= 124 (15.5%) | Isolated hirsutism<br>n= 73 (9.1%) | PCOS<br>n= 55 (6.9%) | p-value |
| --- | --- | --- | --- | --- | --- |
| Age, years | 18.0 (15.0 – 27.0) | 22.0 (17.0 – 30.0) | 17.0 (15.0 – 22.0) | 21.0 (17.0 – 25.0) | **0.007** |
| Adolescent | 306 (56.3) | 46 (37.1) | 46 (63.0) | 25 (45.5) | **<0.001** |
| Adult | 238 (43.8) | 78 (62.9) | 27 (37.0) | 30 (54.5) |  |
| Residence |  |  |  |  |  |
| Urban | 250 (46.0) | 64 (51.6) | 24 (32.9) | 34 (61.8) | **0.007** |
| Rural | 294 (54.0) | 60 (48.4) | 49 (67.1) | 21 (38.2) |  |
| Division |  |  |  |  |  |
| Dhaka 9.6% | 30 (5.5) | 10 (8.1) | 7 (9.6) | 5 (9.1) | **<0.001** |
| Chattagram 2.7% | 57 (10.5) | 14 (11.3) | 1 (1.4) | 2 (3.6) |  |
| Rajshahi 12.5% | 56 (10.3) | 13 (10.5) | 15 (20.5) | 12 (21.8) |  |
| Khulna 2.2% | 66 (12.1) | 18 (14.5) | 4 (5.5) | 2 (3.6) |  |
| Barishal 7.6% | 69 (12.7) | 14 (11.3) | 2 (2.7) | 7 (12.7) |  |
| Sylhet 7.7% | 103 (18.9) | 20 (16.1) | 20 (27.4) | 12 (21.8) |  |
| Rangpur, 7.9% | 53 (9.7) | 20 (16.1) | 20 (27.4) | 8 (14.5) |  |
| Mymensingh, 5.2% | 110 (20.2) | 15 (12.1) | 4 (5.5) | 7 (12.7) |  |
| Educational status |  |  |  |  |  |
| <SSC | 220 (40.4) | 58 (46.8) | 26 (35.6) | 15 (27.3) | 0.307 |
| SSC-HSC | 260 (47.8) | 52 (41.9) | 39 (53.4) | 33 (60.0) |  |
| >HSC | 64 (11.8) | 14 (11.3) | 8 (11.0) | 7 (12.7) |  |
| Occupation |  |  |  |  |  |
| Student | 357 (65.6) | 60 (48.4) | 57 (78.1) | 34 (61.8) | **0.002** |
| Housewife | 157 (28.9) | 55 (44.4) | 14 (19.2) | 19 (34.5) |  |
| Others | 30 (5.5) | 9 (7.3) | 2 (2.7) | 2 (3.6) |  |
| Socio-economic status |  |  |  |  |  |
| Lower | 180 (33.1) | 44 (35.5) | 24 (32.9) | 17 (30.9) | 0.920 |
| Middle | 178 (32.7) | 44 (35.5) | 23 (31.5) | 21 (38.2) |  |
| Upper | 186 (34.2) | 36 (29.0) | 26 (35.6) | 17 (30.9) |  |
| Religion |  |  |  |  |  |
| Islam | 488 (89.7) | 115 (92.7) | 65 (89.0) | 50 (90.9) | 0.753 |
| Sanatan | 56 (10.3) | 9 (7.3) | 8 (11.0) | 5 (9.1) |  |

Within parentheses are percentages over the column total for qualitative variables
Data were expressed in frequency (%) for qualitative and median (IQR) for quantitative variables
Chi-squared or Kruskal-Wallis test was done as appropriate

**Table 2:** Personal history, family history, and lifestyle factors of the study population (n= 796)

| Variables | Control<br>n= 544 | Isolated irregular<br>cycle, n= 124 | Isolated hirsutism<br>n= 73 | PCOS<br>n= 55 | p-value |
| --- | --- | --- | --- | --- | --- |
| Age of menarche, years | 13.0 (12.0 – 13.0) | 13.0 (12.0 – 14.0) | 12.0 (12.0 – 14.0) | 12.0 (12.0 – 14.0) | 0.295 |
| Gynecological age | 5.0 (3.0, 14.0) | 9.5 (4.0, 17.0) | 5.0 (2.5, 10.0) | 8.0 (5.0, 13.0) | **<0.001** |
| ≥3 years | 415 (76.3) | 105 (84.7) | 55 (75.3) | 49 (89.1) | **0.038** |
| ≥8 years | 360 (66.2) | 103 (83.1) | 46 (63.0) | 44 (80.0) | **<0.001** |
| Personal history |  |  |  |  |  |
| Marital status |  |  |  |  |  |
| Unmarried | 358 (65.8) | 56 (47.5) | 56 (71.8) | 33 (60.0) | **0.001** |
| Married & others | 186 (34.2) | 62 (52.5) | 22 (28.2) | 22 (40.0) |  |
| Subfertility, n= 308 | 32/197 (16.2) | 24/65 (36.9) | 4/23 (17.4) | 9/23 (39.1) |  |
| GDM, n= 240 | 5/153 (3.3) | 1/51 (2.0) | 0/21 (0.0) | 0/15 (0.0) |  |
| Family history (1<sup>st</sup> degree relatives) |  |  |  |  |  |
| Irregular cycle | 72 (13.2) | 40 (32.3) | 17 (23.3) | 24 (43.6) | **<0.001** |
| Hirsutism | 28 (5.1) | 13 (10.5) | 12 (16.4) | 12 (21.8) | **<0.001** |
| DM | 145 (26.7) | 33 (26.6) | 14 (19.2) | 14 (25.5) | 0.589 |
| Diet |  |  |  |  |  |
| Fruits ≥2 servings/day | 77 (14.2) | 18 (15.3) | 13 (16.7) | 10 (18.2) | 0.751 |
| Vegetables ≥2 servings/day | 227 (41.7) | 51 (41.1) | 27 (37.0) | 16 (29.1) | 0.296 |
| Outside meals ≥3 /week | 41 (7.5) | 12 (9.7) | 6 (8.2) | 6 (10.9) | 0.749 |
| Fast food/ oily snacks ≥3 /week | 197 (36.2) | 46 (37.1) | 27 (37.0) | 15 (27.3) | 0.590 |
| Smokeless tobacco consumption | 15 (3.0) | 6 (5.1) | 4 (5.5) | 1 (1.9) |  |
| Physical activity level | 15 (3.0) | 6 (5.1) | 4 (5.5) | 1 (1.9) |  |
| Low <600 MET/ week | 207 (38.1) | 37 (29.8) | 27 (37.0) | 19 (34.5) |  |
| Moderate 6000 – 2999 MET/week | 285 (52.4) | 70 (56.5) | 39 (53.4) | 30 (54.5) | 0.676 |
| Vigorous >2999 MET/week | 52 (9.6) | 17 (13.7) | 7 (9.6) | 6 (10.9) |  |
| Sitting time ≥6 hours/ day | 118 (21.7) | 27 (21.8) | 11 (15.1) | 9 (16.4) | 0.484 |
| Screen time ≥2 hours/day | 207 (38.1) | 34 (27.4) | 26 (35.6) | 26 (47.3) | 0.053 |
| Sleep |  |  |  |  |  |
| Bedtime after 10 pm | 307 (56.5) | 71 (57.3) | 41 (56.2) | 43 (78.2) | **0.020** |
| Wake up after 7 am | 144 (26.5) | 33 (26.6) | 23 (31.5) | 22 (40.0) | 0.162 |
| Adequate sleep duration* | 372 (68.4) | 84 (67.7) | 53 (72.6) | 30 (54.5) | 0.153 |

Metabolic equivalent task (MET); \*Adequate sleep duration: Adolescents: 8 – 10 hours, Adults: 7 – 9 hours
Within parentheses are percentages over the column total for qualitative variables
Data were expressed in frequency (%) for qualitative and median (IQR) for quantitative variables
Chi-squared or Kruskal-Wallis test was done as appropriate

There were significant differences in gynecological age groups (χ² = 17.9, φc = 0.2) and marital status (χ² = 24.9, φc = 0.2) among the study groups. Those with IIC had a higher frequency of married and others (ASR=+4.5). Those with IIC had a higher gynecological age than those with ISH (p=0.003) and the control women (p=0.002). Family history (FH) of irregular cycles (χ² = 48.0, φc = 0.3) and hirsutism (χ² = 27.8, φc = 0.2) differed significantly among the study groups. The frequency of a FH of irregular cycles was substantially higher in women with PCOS (ASR=+4.8) and IIC (ASR=+4.0), but lower in controls (ASR=-6.3). The frequency of FH of hirsutism was higher among women with PCOS (ASR=+3.8), but lower in controls (ASR=-4.6). Among various lifestyle factors, only the frequency of women who slept after 10 pm was significantly higher among women with PCOS (χ^2^ = 9.9, ASR = +3.1, φc = 0.1) (Table 2).

There were significant differences in the frequencies of BMI-status (χ^2^=12.4, φ_c_=0.2), WC-status χ =17.5, φ_c_=0.2), AN (χ =16.0, φ_c_=0.1), acne (χ =49.7, φ_c_=0.3), goitre (χ =15.6, φ_c_=0.1), and metabolic syndrome (χ^2^=8.2, φ_c_=0.1) among the study groups. Nearly 42% of women with PCOS had general and central obesity, one-third had abnormal glycemic status, and 11% had possible MASH. The frequency of AN was higher in the PCOS group (χ2 = 16.0, ASR = +3.9).

The frequency of goitre (χ^2^=15.6) was higher in women with IIC (ASR = +3.4). The frequency of acne (χ^2^=49.7) was higher in those with ISH (ASR = +5.2) and PCOS (ASR = +4.3). The mFG score progressively decreased from ISH to PCOS, then IIC, and then to controls. (Table 3).

**Table 3:**
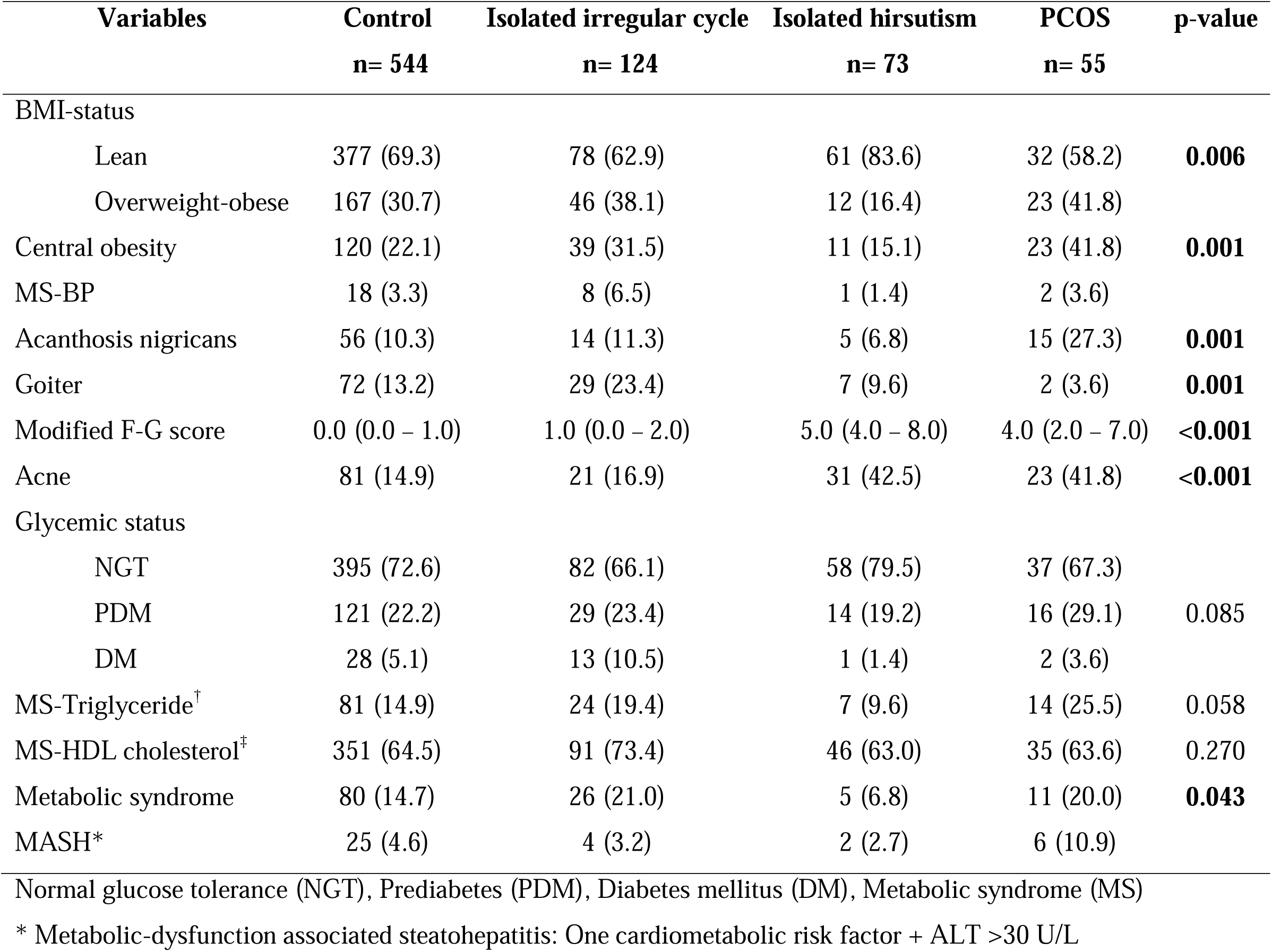
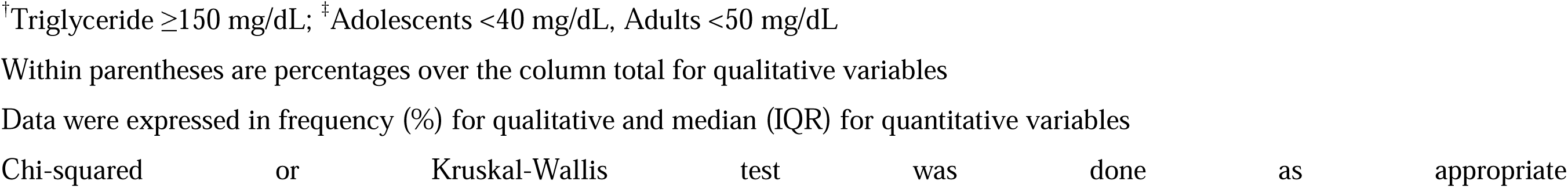
Physical examination and investigation findings of the study population (n= 796)

| Variables | Control<br>n= 544 | Isolated irregular cycle<br>n= 124 | Isolated hirsutism<br>n= 73 | PCOS<br>n= 55 | p-value |
| --- | --- | --- | --- | --- | --- |
| BMI-status |  |  |  |  |  |
| Lean | 377 (69.3) | 78 (62.9) | 61 (83.6) | 32 (58.2) | **0.006** |
| Overweight-obese | 167 (30.7) | 46 (38.1) | 12 (16.4) | 23 (41.8) |  |
| Central obesity | 120 (22.1) | 39 (31.5) | 11 (15.1) | 23 (41.8) | **0.001** |
| MS-BP | 18 (3.3) | 8 (6.5) | 1 (1.4) | 2 (3.6) |  |
| Acanthosis nigricans | 56 (10.3) | 14 (11.3) | 5 (6.8) | 15 (27.3) | **0.001** |
| Goiter | 72 (13.2) | 29 (23.4) | 7 (9.6) | 2 (3.6) | **0.001** |
| Modified F-G score | 0.0 (0.0 – 1.0) | 1.0 (0.0 – 2.0) | 5.0 (4.0 – 8.0) | 4.0 (2.0 – 7.0) | **<0.001** |
| Acne | 81 (14.9) | 21 (16.9) | 31 (42.5) | 23 (41.8) | **<0.001** |
| Glycemic status |  |  |  |  |  |
| NGT | 395 (72.6) | 82 (66.1) | 58 (79.5) | 37 (67.3) |  |
| PDM | 121 (22.2) | 29 (23.4) | 14 (19.2) | 16 (29.1) | 0.085 |
| DM | 28 (5.1) | 13 (10.5) | 1 (1.4) | 2 (3.6) |  |
| MS-Triglyceride<sup>†</sup> | 81 (14.9) | 24 (19.4) | 7 (9.6) | 14 (25.5) | 0.058 |
| MS-HDL cholesterol<sup>‡</sup> | 351 (64.5) | 91 (73.4) | 46 (63.0) | 35 (63.6) | 0.270 |
| Metabolic syndrome | 80 (14.7) | 26 (21.0) | 5 (6.8) | 11 (20.0) | **0.043** |
| MASH* | 25 (4.6) | 4 (3.2) | 2 (2.7) | 6 (10.9) |  |

Normal glucose tolerance (NGT), Prediabetes (PDM), Diabetes mellitus (DM), Metabolic syndrome (MS)
\* Metabolic-dysfunction associated steatohepatitis: One cardiometabolic risk factor + ALT >30 U/L
<sup>†</sup>Triglyceride $\geq 150$ mg/dL; <sup>‡</sup>Adolescents $< 40$ mg/dL, Adults $< 50$ mg/dL
Within parentheses are percentages over the column total for qualitative variables
Data were expressed in frequency (%) for qualitative and median (IQR) for quantitative variables
Chi-squared or Kruskal-Wallis test was done as appropriate

### Adult vs. adolescent PCOS

The comparison between adolescent and adult PCOS was depicted in Table 4 and Table 5. There were significant differences in occupation (χ^2^=17.7; Phi, φ_c_=-0.6), marital status (χ^2^=15, φ_c_=0.5), and gynecological age (χ^2^=16.5, φ_c_=0.6) (Table 4). Adolescents with PCOS were physically less active (χ² = 13.1, φc = −0.5) and spent more time sitting (χ² = 4.5, φc = −0.3) than adults with PCOS. However, the frequencies of low HDL-cholesterol (χ^2^ = 11.1, φc = 0.5) and MetS (χ^2^ = 7.3, φc = 0.4) were higher among adults with PCOS than among adolescents (Table 5).

**Table 4:** Baseline characteristics between adolescent and adult PCOS, n=55.

| Variables | Adolescent PCOS<br>n= 25 | Adult PCOS<br>n= 30 | p-value |
| --- | --- | --- | --- |
| Residence |  |  |  |
| Urban | 16 (64.0) | 18 (60.0) | 0.761 |
| Rural | 9 (36.0) | 12 (40.0) |  |
| Educational status |  |  |  |
| <SSC | 9 (36.0) | 6 (20.0) | 0.091 |
| SSC & above | 16 (64.0) | 24 (80.0) |  |
| Occupation |  |  |  |
| Student | 23 (92.0) | 11 (42.3) | **<0.001** |
| Housewife & others | 2 (8.0) | 15 (57.7) |  |
| Socio-economic status |  |  |  |
| Lower | 9 (36.0) | 8 (26.7) |  |
| Middle | 7 (28.0) | 14 (46.7) | 0.365 |
| Upper | 9 (36.0) | 8 (26.7) |  |
| Religion |  |  |  |
| Islam | 23 (92.0) | 27 (90.0) | 1.00 |
| Sanatan | 2 (8.0) | 3 (10.0) |  |
| Marital status |  |  |  |
| Unmarried | 22 (88.0) | 11 (36.7) | **<0.001** |
| Married & others | 3 (12.0) | 19 (63.3) |  |
| Age of menarche, years | 12.0 (11.0 – 14.0) | 13.0 (12.0- 14.0) | 0.119 |
| Gynecological age | 5.0 (2.5 – 6.0) | 12.0 (9.0 – 20.3) | **<0.001** |
| ≥3 years | 19 (76.0) | 30 (100.0) | **0.006** |
| Family history |  |  |  |
| Irregular cycle | 12 (48.0) | 12 (40.0) | 0.551 |
| Hirsutism | 7 (28.0) | 5 (16.7) | 0.311 |
| DM | 6 (24.0) | 8 (27.6) | 0.764 |

Data were expressed in frequency (%) or median (IQR); Within parentheses are percentages over the column total for qualitative variables; Chi-squared/Fisher's exact test or Mann-Whitney U test was done as appropriate

**Table 5:** Lifestyle factors, physical examination findings, and investigation profile between adolescent and adult PCOS, n=55.

| Variables | Adolescent PCOS | Adult PCOS | p-value |
| --- | --- | --- | --- |
| Diet |  |  |  |
| Fruits $\geq 2$ servings/day | 4 (16.0) | 6 (20.0) | 0.741 |
| Vegetables $\geq 2$ servings/day | 5 (20.0) | 11 (36.7) | 0.175 |
| Outside meals $\geq 3$ /week | 3 (12.0) | 3 (10.0) | 1.000 |
| Fast food/ oily snacks $\geq 3$ /week | 10 (40.0) | 5 (16.7) | 0.053 |
| Physical activity level |  |  |  |
| Low <600 MET/ week | 15 (60.0) | 4 (13.3) | **<0.001** |
| Moderate- vigorous $\geq 600$ MET/week | 10 (40.0) | 26 (86.7) | |
| Sitting time $\geq 6$ hours/ day | 7 (28.0) | 2 (6.7) | **0.033** |
| Screen time $\geq 2$ hours/day | 9 (36.0) | 7 (23.3) | 0.303 |
| Sleep |  |  |  |
| Bedtime after 10 pm | 18 (72.0) | 25 (83.3) | 0.311 |
| Wake up after 7 am | 13 (52.0) | 9 (30.0) | 0.097 |
| Inappropriate sleep duration* | 13 (52.0) | 12 (40.0) | 0.373 |
| BMI-status |  |  |  |
| Lean | 16 (64.0) | 16 (53.3) | 0.425 |
| Overweight-obese | 9 (36.0) | 14 (46.7) |  |
| Central obesity | 7 (28.0) | 16 (53.3) | 0.058 |
| MS-BP | 0 (0.0) | 2 (6.7) | 0.495 |
| Acanthosis nigricans | 6 (24.0) | 9 (30.0) | 0.619 |
| Goiter | 2 (8.0) | 0 (0.0) | 0.202 |
| Modified F-G score | 5.0 (3.5 – 7.5) | 4.0 (1.5 – 7.3) | 0.441 |
| Acne | 13 (52.0) | 10 (33.3) | 0.162 |
| Glycemic status |  |  |  |
| NGT | 18 (72.0) | 18 (69.2) | 0.828 |
| AGT | 7 (28.0) | 8 (30.0) |  |
| MS-Triglyceride | 5 (20.0) | 9 (30.0) | 0.397 |
| MS-HDL cholesterol | 10 (40.0) | 25 (83.3) | **0.001** |
| Metabolic syndrome | 1 (4.0) | 10 (33.3) | **0.007** |
| MASH<sup>†</sup> | 2 (8.0) | 4 (13.3) | 0.678 |

\*Adequate sleep duration: Adolescents: 8 – 10 hours, Adults: 7 – 9 hours
<sup>†</sup>Metabolic-dysfunction associated steatohepatitis: One cardiometabolic risk factor + ALT >30 U/L
Data were expressed in frequency (%) or median (IQR)
Within parentheses are percentages over the column total for qualitative variables
Chi-squared/Fisher's exact test or Mann-Whitney U test was done as appropriate

### Characteristics within the adolescent and adult participants

Due to the small number of participants, we could not compare the four study groups within adolescents and adults for all the variables (Supplementary Table 1, Supp. Table 2, Supp. Table 3, Supp. Table 4, Suppl. Table 5, and Suppl. Table 6).

### Characteristics of the adolescent study groups

Among the adolescent group, the women with PCOS were older than the control group (post hoc p=0.043). The mFGS was higher in women with ISH than in all other groups. The mFGS of women with PCOS was higher than the rest of the other two groups. The IIC and control groups had similar mFGS. The PCOS group had higher residence in urban areas than rural areas χ =11.9, φ_c_=+3, p=0.008). Significant differences in AN (χ =9.0, φ_c_=0.2, p=0.030), wake up after 7 am (χ^2^=10.1, φ_c_=0.2, p=0.018), and screen time ≥2 hours/day (χ^2^=8.4, φ_c_=0.1, p=0.038) were observed across the four groups. Higher frequency of acne (χ^2^=35.2, p<0.001) in ISH (ASR=+3.6) and PCOS (ASH=+4.3), but lower in the control group (ASR=-3.8), was observed. Family history of irregular cycle (χ^2^=24.7, φ_c_=0.2, p<0.001) and hirsutism (χ^2^=22.2, φ_c_=0.3, p<0.001) were higher in the PCOS group (ASR= +4.3 for both) and lower in the control group (ASR=-3.9 & −3.4, respectively). Women with PCOS (ASR=+3.7) had a higher frequency of overweight-obesity (χ^2^=16.0, φ_c_=0.2, p=0.001).

### Characteristics of the adult study groups

Within adults, significant differences in goiter (χ^2^=9.1, φ_c_=0.2, p=0.028), marital status (χ^2^=11.4, φ_c_=0.2, p=0.010), occupation (χ^2^=8.3, φ_c_=0.2, p=0.039), family history of hirsutism (χ^2^=13.3, φ_c_=0.2, p=0.004), and glycemic status (χ^2^=9.8, φ_c_=0.2, p=0.020) were observed across the groups. Acne (χ^2^=20.2, φ_c_=0.2, p<0.001) was higher in the ISH group (ASR=+3.8) and lower in the control group (ASR=-3.2). Family history of irregular cycles was higher (χ^2^=23.6, φ_c_=0.3, p<0.001) in the IIC (ASR=+3.3) and lower in controls (ASR=-4.8). The mFG scores were similar between PCOS and ISH. Both PCOS and ISH had higher mFG scores than IIC and controls. The IIC group had higher mFG scores than the control group.

## Discussion

The current study found a 6.9% prevalence of PCOS among reproductive-aged Bangladeshi women using the International Evidence-based Guidelines for PCOS, 2023; more specifically, by the stepwise decision-making and AMH. The prevalence was 5.9% and 8.0%, respectively, in adolescents and adults. IIC were present in 15.5%, ISH was present in 9.1%, and those without irregular cycles and significant hirsutism (controls) were 68.4%. The FH of irregular cycles and hirsutism was significantly higher among women with PCOS than in controls. Late bedtime was more frequent among them. Besides, the frequencies of AN, acne, and mFG score were higher among women with PCOS than in controls. Those with IIC had higher frequencies of adults, married, housewives, and a FH of irregular cycles, goitre, and a higher mFG score than in controls. Those with ISH had a lower age and gynecological age than those with IIC. Additionally, the women in the ISH group had the highest frequency of acne and a higher mFG score. Adolescents with PCOS were physically more inactive and spent more time sitting, but had lower low-HDL cholesterol and metabolic syndrome than adults with PCOS.

### Prevalence of PCOS

A recent meta-analysis revealed a nearly 10% global prevalence of PCOS, although reported figures vary widely across individual studies, ranging from 5% to 21% (Salari N, et al., 2024). This variability is influenced by numerous factors, including geographical region, racial and ethnic differences, the specific diagnostic criteria employed, the age demographic of the study population, and the nature of the cohort (e.g., community-based versus clinical samples) (Yasmin A, et al., 2022). In South Asia, women are recognized as being more predisposed to PCOS. For instance, a study in a U.S. cohort found the prevalence among South Asian women to be 3.3%, which was 2.6-fold higher than that among Chinese women. Regional studies showed the prevalence of PCOS is 6.3% in Sri Lanka (15 – 39 years, 2005-06) and 7.2 – 19.6% in India (18 – 40 years, 2018-22), depending on the criteria used. As we used a step-wise decision-making process to diagnose our cases, our findings are not entirely comparable to those studies. However, the prevalence found in our research is at the lower end of the published ranges. Notably, our participants’ lower age range is 10 years, and we have more participants from the adolescent age group. Additionally, we used the PCOM criteria from the age of 20. Moreover, those presented with isolated irregular cycles and isolated significant hirsutism (collectively ∼25%) might be mild cases of PCOS requiring further investigations, such as free testosterone, sex-hormone binding globulin, androstenedione, dehydroepiandrosterone sulphate, follicular phase progesterone, etc. Besides, ultrasonography might detect more cases of PCOM than AMH. In a meta-analysis, the prevalence of PCOS among adolescents was reported at 6.3%, slightly higher than our finding. Along with the described factors, many of the studies included a selective population rather than community participants. Besides more stringent criteria in adolescents, it is also plausible that the syndrome naturally progresses over time, with more women meeting the whole constellation of diagnostic criteria as they advance through their reproductive years, contributing to a higher prevalence in adults.

The significantly higher prevalence of a FH of irregular cycles and hirsutism among women with PCOS compared to controls provides further epidemiological support for the well-established genetic and familial predisposition to PCOS (Goodarzi et al., 2011). This reinforces the need to incorporate FH into clinical assessment for risk stratification.

The association of late bedtime with PCOS in this cohort adds to the growing body of evidence implicating circadian rhythm disruption in PCOS pathogenesis. Late sleep patterns may exacerbate hormonal imbalances and metabolic dysregulation, including insulin resistance, which are key components of the syndrome (Moghetti & Tosi, 2021). This finding suggests that sleep hygiene may be a potentially modifiable lifestyle factor for intervention.

As expected, cutaneous markers of hyperandrogenism (significant hirsutism and acne) and insulin resistance (acanthosis nigricans) were significantly more prevalent in the PCOS group compared to the controls (Escobar-Morreale, 2018). This underscores the systemic nature of PCOS, where hyperandrogenism and metabolic dysfunction manifest visibly on the skin, extending the impact of the syndrome beyond gynecological and reproductive health.

Despite following unhealthy lifestyles, the adolescent PCOS group had fewer metabolic abnormalities, suggesting metabolic complications, particularly insulin resistance and dyslipidemia, tend to develop and worsen over time in PCOS (Moran et al., 2010). The adolescents may use more cell phones, the internet, and social media than adults, and perform fewer physical activities, as the prevalence of PCOS was found to be higher in urban areas where places for physical activities are limited. This underscores the critical window of opportunity during adolescence for implementing lifestyle interventions that focus on improving physical activity and diet to mitigate the future progression of significant cardiometabolic diseases.

### Participants with an isolated diagnostic feature

A key insight from this study is that a significant proportion of women present with isolated PCOS features: 15. 5% with IIC and 9. 9.1% with ISH. This high prevalence of isolated symptoms, compared to the 6.9% meeting full diagnostic criteria, underscores the considerable heterogeneity of PCOS manifestations. It highlights the importance of distinguishing between the full syndrome and isolated symptoms in clinical practice. Overdiagnosis based on isolated features can cause unnecessary anxiety and interventions, while underdiagnosis of evolving PCOS may delay management, especially regarding metabolic risks. This distinction is crucial, particularly in settings like Bangladesh, where social stigma related to features such as hirsutism and the risk of overlooking metabolic consequences has significant psychosocial and health impacts (Hasan M, 2022). These findings suggest a large group of women who may be’ at-risk” or exhibit a milder phenotype, needing monitoring rather than immediate diagnosis. Women with IIC were generally older, married, housewives, with a stronger family history of irregular cycles and goiter, and higher mFG scores than controls, potentially representing a subset where ovulatory dysfunction is a primary and possibly chronic feature. The current guidelines define any cycle longer than three months as abnormal. This expands case detection and indicates that regular cycles do not necessarily exclude irregular cycles, as menstrual patterns in women with PCOS are highly variable. The link with goiter is intriguing and suggests a possible connection with thyroid dysfunction or autoimmunity, which warrants further investigation (P HH et al., 2024). This group may include women with evolving PCOS, where hyperandrogenic features are subclinical or emerging, or potentially a distinct phenotype prone to chronic anovulation. The younger age and lower gynecological age of women with ISH, along with the higher prevalence of acne and elevated mFG scores, strongly suggest that hyperandrogenic features, particularly hirsutism, can be an early sign in PCOS development, possibly appearing before notable menstrual irregularities (Lizneva et al., 2016). This emphasizes the importance of thoroughly evaluating hyperandrogenism even in younger women with regular cycles.

The stepwise decision-making may reduce the number of investigations; however, it may increase the number of physician visits. We have to use the same serum several times to reach the diagnosis, which may not be possible in clinical settings.

### Limitations

Due to the stepwise decision-making process, we were unable to comment on the phenotypes of PCOS. Similarly, further investigations could identify additional women with PCOS from the IIC or ISH group. Because there was only one period of sample collection, we could not exclude non-classic congenital adrenal hyperplasia and hypogonadism, which require follicular phase sample collection. Additionally, we measured TT and AMH in only 100 controls to establish their cut-offs, which may not be the true representative of the population-specific values.

## Conclusions

A stepwise decision-making process and the use of AMH found a 6.9% prevalence of PCOS, indicating a significant health burden among Bangladeshi women of reproductive age. This study enhances the understanding of PCOS epidemiology in South Asia by illustrating both its prevalence and phenotypic variation among Bangladeshi women. The findings emphasize the need for age-specific and symptom-specific assessment strategies and suggest potential genetic and lifestyle factors in the development of PCOS. Further longitudinal studies are necessary to track the progression of isolated symptoms to full-blown PCOS and to create targeted interventions.

## Data Availability

All data produced in the present work are contained in the manuscript

## Declarations

Funding statement: The Fund was arranged by Prof. M A Hasanat, Endocrinology, BMU Conflict of interest: Nil

## Acknowledgement

Dr. Debasish Kumar Ghosh, Assistant Professor of Endocrinology, Khulna Medical College; Dr. Mohammad Atiqur Rahman, Assistant Professor of Endocrinology, National Institute of Neurosciences, Dhaka; Dr. Abu Jar Gaffar, Naogaon Medical College; Dr. Md. Tozammel Haque, Senior Consultant (Medicine), 250 Bedded General Hospital, Thakurgaon; Dr. Sarwar Rahman Tusher, Consultant, 250 Bedded General Hospital, Thakurgaon; Dr. Md Abdullah-Al-Maruf, Consultant (Medicine), Moulovibazar General Hospital; Dr. Farhana Qayum, Consultant (Dermatologist), BRB Hospital Ltd.; Dr. Md. Kamrul Azad, Consultant, Government Employee Hospital, Dhaka; Dr. Md. Abdullah Al Mamun, FCPS part 2 trainee, BMU

---

**Supplementary Table 1:** Baseline characteristics of the adolescents, n= 423.

| Variables | Control<br>n= 306 (72.3%) | Isolated irregular cycle<br>n= 46 (10.9%) | Isolated hirsutism<br>n= 46 (10.9%) | PCOS<br>n= 25 (5.9%) | p-value |
| --- | --- | --- | --- | --- | --- |
| Age, years | 15.0 (14.0 – 17.0) | 16.0 (14.0 – 17.3) | 16.0 (15.0 – 17.0) | 17.0 (16.0 – 18.0) | **0.046** |
| Residence |  |  |  |  |  |
| Urban | 106 (34.6) | 18 (39.1) | 11 (23.9) | 16 (64.0) (+3) | **0.008** |
| Rural | 200 (65.4) | 28 (60.9) | 35 (76.1) | 9 (36.0) (-3) |  |
| Division |  |  |  |  |  |
| Dhaka | 7 (2.3) | 3 (6.5) | 4 (8.7) | 1 (4.0) |  |
| Chattagram | 24 (7.8) | 4 (8.7) | 1 (2.2) | 1 (4.0) |  |
| Rajshahi | 23 (7.5) | 1 (2.2) | 9 (19.6) | 4 (16.0) |  |
| Khulna | 37 (12.1) | 7 (15.2) | 2 (4.3) | 1 (4.0) |  |
| Barishal | 21 (6.9) | 3 (6.5) | 1 (2.2) | 1 (4.0) |  |
| Sylhet | 84 (27.5) | 13 (28.3) | 17 (37.0) | 11 (44.0) |  |
| Rangpur | 39 (12.7) | 12 (26.1) | 9 (19.6) | 4 (16.0) |  |
| Mymensingh | 71 (23.2) | 3 (6.5) | 3 (6.5) | 2 (8.0) |  |
| Educational status |  |  |  |  |  |
| <SSC | 140 (45.8) | 23 (50.0) | 18 (39.1) | 9 (36.0) | 0.574 |
| ≥SSC | 166 (54.2) | 23 (50.0) | 28 (60.9) | 16 (64.0) |  |
| Occupation |  |  |  |  |  |
| Student | 294 (96.1) | 43 (93.5) | 44 (95.7) | 23 (92.0) |  |
| Housewife & others | 12 (3.9) | 3 (6.5) | 2 (4.3) | 2 (8.0) |  |

| Socio-economic status |  |  |  |  |  |
| --- | --- | --- | --- | --- | --- |
| Lower | 106 (34.6) | 14 (30.4) | 14 (30.4) | 9 (36.0) | 0.918 |
| Middle | 104 (34.0) | 16 (34.8) | 15 (32.6) | 6 (24.0) |  |
| Upper | 96 (31.4) | 16 (34.8) | 17 (37.0) | 10 (40.0) |  |
| Religion |  |  |  |  |  |
| Islam | 270 (88.2) | 43 (93.5) | 40 (87.0) | 23 (92.0) | 0.675 |
| Sanatan | 36 (11.8) | 3 (6.5) | 6 (13.0) | 2 (8.0) |  |

Within parentheses are percentages over the column total for qualitative variables
Data were expressed in frequency (%) for qualitative and median (IQR) for quantitative variables
Chi-squared or Kruskal-Wallis test was done as appropriate

**Supplementary Table 2:** Personal and family history of the adolescents, n= 423.

| Variables | Control<br>n= 306 | Isolated irregular<br>cycle, n=46 | Isolated hirsutism<br>n= 46 | PCOS<br>n= 25 | p-value |
| --- | --- | --- | --- | --- | --- |
| Age of menarche, years | 12.0 (12.0 – 13.0) | 12.0 (12.0 – 13.0) | 12.0 (12.0 – 13.0) | 12.0 (11.0 – 14.0) | 0.897 |
| Gynecological age | 3.0 (2.0 – 4.0) | 4.0 (1.8 – 4.0) | 3.0 (2.0 – 5.0) | 5.0 (2.5 – 6.0) | 0.072 |
| ≥3 years | 123 (40.2) | 25 (54.3) | 19 (41.3) | 14 (56.0) | 0.159 |
| Personal history |  |  |  |  |  |
| Marital status |  |  |  |  |  |
| Unmarried | 299 (97.7) | 44 (95.7) | 44 (95.7) | 22 (88.0) |  |
| Married & others | 7 (2.3) | 2 (4.3) | 2 (4.3) | 3 (12.0) |  |
| Subfertility, n= 21 | 1/10 | 1/4 | 0/4 | 2/3 |  |
| Family history (1<sup>st</sup> degree relatives) |  |  |  |  |  |
| Irregular cycle | 38 (12.4) (-3.9) | 12 (26.1) | 9 (19.6) | 12 (48.0) (4.3) | **<0.001** |
| Hirsutism | 13 (4.2) (-3.4) | 4 (8.7) | 5 (10.9) | 7 (28.0) (4.3) | **<0.001** |
| DM | 66 (21.6) | 13 (28.3) | 5 (10.9) | 6 (24.0) | 0.218 |
| Diet |  |  |  |  |  |
| Fruits ≥2 serving/day | 10 (3.3) | 2 (4.3) | 0 (0.0) | 1 (4.0) |  |
| Vegetables ≥2 serving/day | 7 (2.3) | 4 (8.7) | 0 (0.0) | 0 (0.0) |  |
| Outside meals ≥3 /week | 14 (4.6) | 1 (2.2) | 4 (8.7) | 1 (4.0) |  |
| Fast food/ oily snacks ≥3 /week | 153 (50.0) | 28 (60.9) | 21 (45.7) | 10 (40.0) | 0.318 |
| Smokeless tobacco consumption | 1 | 0 | 0 | 0 |  |
| Physical activity level |  |  |  |  |  |
| Low <600 MET/ week | 160 (52.3) | 26 (56.5) | 20 (43.5) | 15 (60.0) | 0.502 |
| Moderate - Vigorous | 146 (47.7) | 20 (43.5) | 26 (56.5) | 10 (40.0) |  |
| Sitting time $\geq 6$ hours/ day | 82 (26.8) | 14 (30.4) | 7 (15.2) | 7 (28.0) | 0.334 |
| Screen time $\geq 2$ hours/day | 113 (36.9) | 12 (26.1) | 15 (32.6) | 15 (60.0) (2.5) | **0.038** |
| Sleep |  |  |  |  |  |
| Bedtime after 10 pm | 152 (49.7) | 27 (58.7) | 26 (56.5) | 18 (72.0) | 0.122 |
| Wake up after 7 am | 77 (25.2) | 13 (28.3) | 17 (37.0) | 13 (52.0) (2.7) | **0.018** |
| Adequate sleep duration* | 212 (69.3) | 32 (69.6) | 35 (76.1) | 12 (48.0) | 0.101 |

Within parentheses are percentages over the column total for qualitative variables
Data were expressed in frequency (%) for qualitative and median (IQR) for quantitative variables
Chi-squared or Kruskal-Wallis test was done as appropriate

**Supplementary Table 3:** Physical and laboratory findings of the adolescents, n= 423.

| Variables | Control<br>n= 306 | Isolated irregular cycle<br>n= 46 | Isolated hirsutism<br>n= 46 | PCOS<br>n= 25 | p-value |
| --- | --- | --- | --- | --- | --- |
| BMI-status |  |  |  |  |  |
| Lean | 271 (88.6) | 40 (87.0) | 44 (95.7) | 16 (64.0) |  |
| Overweight-obese | 35 (11.4) | 6 (13.0) | 2 (4.3) | 9 (36.0) (3.7) | **0.001** |
| Central obesity | 16 (5.2) | 3 (6.5) | 3 (6.5) | 7 (28.0) |  |
| MS-BP | 1 (0.3) | 0 | 0 | 0 |  |
| Acanthosis nigricans | 24 (7.8) | 5 (10.9) | 2 (4.3) | 6 (24.0) (2.8) |  |
| Goiter | 30 (9.8) | 10 (21.7) | 3 (6.5) | 2 (8.0) |  |
| Modified F-G score | 0.0 (0.0 – 1.0) | 1.0 (0.0 – 2.0) | 5.0 (4.0 – 8.0) | 5.0 (3.5 – 7.5) | **<0.001** |
| Acne | 45 (14.7) (-3.8) | 5 (10.9) | 18 (39.1) (3.6) | 13 (52.0) (4.3) | **<0.001** |
| Glycemic status |  |  |  |  |  |
| NGT | 225 (73.5) | 36 (78.3) | 34 (73.9) | 18 (72.0) | 0.914 |
| AGT | 81 (26.5) | 10 (21.7) | 12 (26.1) | 7 (28.0) |  |
| MS-Triglyceride | 41 (13.4) | 5 (10.9) | 5 (10.9) | 5 (20.0) | 0.694 |
| MS-HDL cholesterol | 173 (56.5) | 28 (60.9) | 25 (54.3) | 10 (40.0) | 0.374 |
| Metabolic syndrome | 13 (4.2) | 2 (4.3) | 2 (4.3) | 1 (4.0) |  |
| MASH* | 7 (2.3) | 0 | 0 | 2 (8.0) |  |

Data were expressed in frequency (%) or median (IQR)
Within parentheses are the percentages over the column total for the qualitative variables

**Supplementary Table 4:**
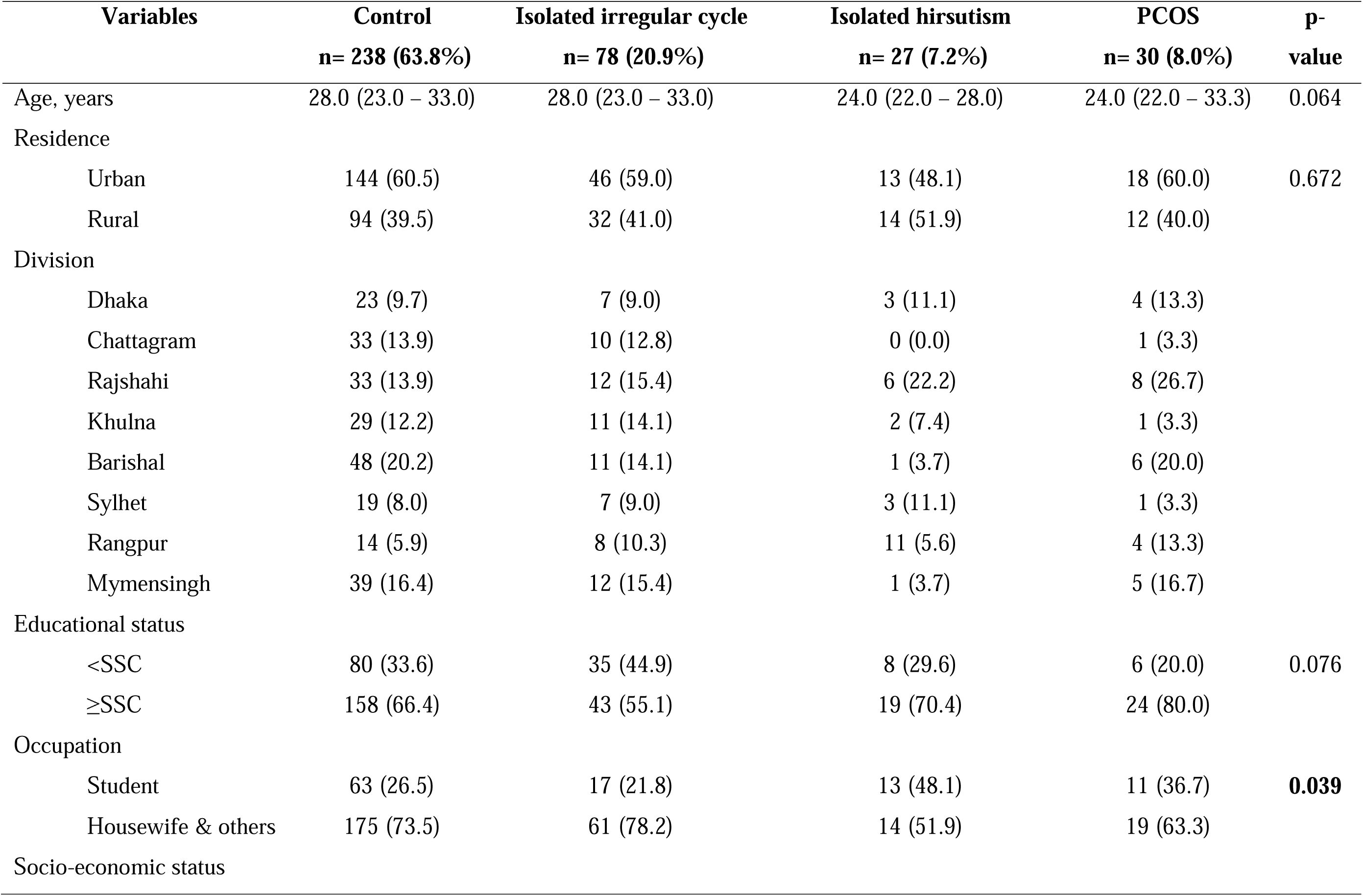
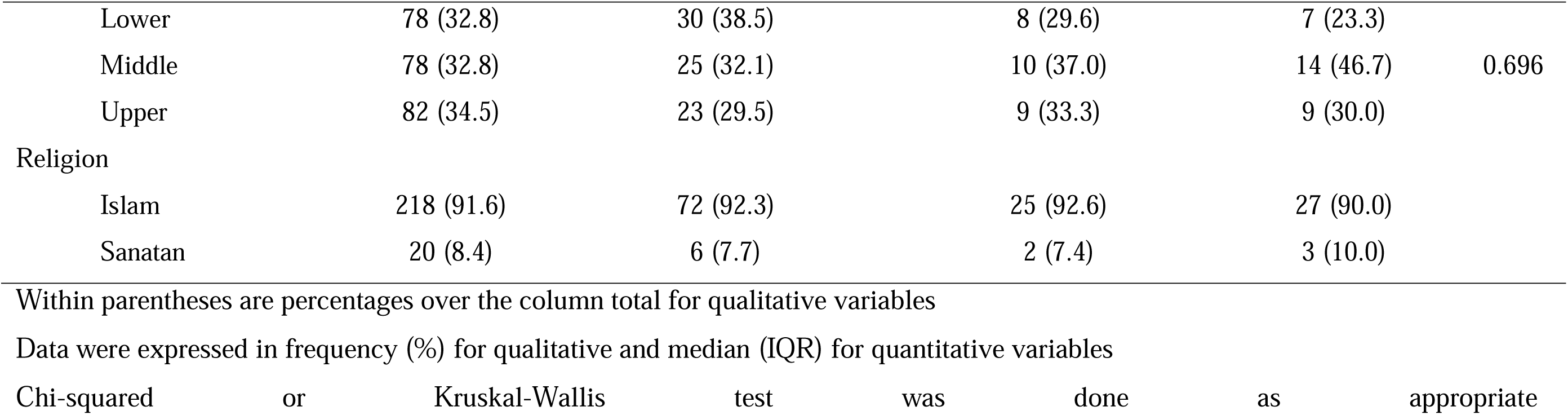
Baseline characteristics of the adult participants, n= 373.

**Supplementary Table 5:** Personal and family history of the adult participants, n= 373.

| Variables | Control<br>n= 238 | Isolated irregular<br>cycle, n= 78 | Isolated hirsutism<br>n= 27 | PCOS<br>n= 30 | p-value |
| --- | --- | --- | --- | --- | --- |
| Age of menarche, years | 13.0 (12.0 – 14.0) | 13.0 (12.0 – 14.0) | 13.0 (12.0 – 14.0) | 13.0 (12.0 – 14.0) | 0.729 |
| Gynecological age | 15.0 (10.0 – 20.0) | 15.0 (11.0 – 19.0) | 11.0 (9.0 – 15.0) | 12.0 (9.0 – 20.3) | 0.059 |
| ≥8 years | 215 (90.3) | 68 (87.2) | 24 (88.9) | 27 (90.0) | 0.886 |
| Personal history |  |  |  |  |  |
| Marital status |  |  |  |  |  |
| Unmarried | 59 (24.8) | 12 (15.4) | 12 (44.4) | 11 (36.7) | **0.010** |
| Married & others | 179 (75.2) | 66 (84.6) | 15 (55.6) | 19 (63.3) |  |
| Subfertility, n= 288 | 31/187 (16.6) | 25/67 (37.3) | 2/14 (14.3) | 7/20 (35.0) |  |
| GDM, n= 228 | 5/148 (3.4) | 1/54 (1.9) | 0/14 | 0/12 |  |
| Family history (1<sup>st</sup> degree relatives) |  |  |  |  |  |
| Irregular cycle 0.3 | 34 (14.3) -4.8 | 28 (35.9) 3.3 | 8 (29.6) | 12 (40.0) | **<0.001** |
| Hirsutism 0.2 | 15 (6.3) | 9 (11.5) | 7 (25.9) 3 | 5 (16.7) | **0.004** |
| DM | 79 (33.2) | 20 (25.6) | 9 (33.3) | 8 (26.7) | 0.591 |
| Diet |  |  |  |  |  |
| Fruits ≥2 servings/day | 3 (1.3) | 4 (5.1) | 0 (0.0) | 1 (3.3) |  |
| Vegetables ≥2 servings/day | 17 (7.1) | 6 (7.7) | 0 (0.0) | 2 (6.7) |  |
| Outside meals ≥3 /week | 9 (3.8) | 4 (5.1) | 1 (3.7) | 3 (10.0) |  |
| Fast food/ oily snacks ≥3 /week | 44 (18.5) | 18 (23.1) | 6 (22.2) | 5 (16.7) | 0.785 |
| Smokeless tobacco consumption |  |  |  |  |  |
| Physical activity level |  |  |  |  |  |
| Low <600 MET/ week | 47 (19.7) | 11 (14.1) | 7 (25.9) | 4 (13.3) | 0.429 |
| Moderate-Vigorous | 191 (80.3) | 67 (85.9) | 20 (74.1) | 26 (86.7) |  |
| Sitting time $\geq 6$ hours/ day | 36 (15.1) | 13 (16.7) | 4 (14.8) | 2 (6.7) | |
| Screen time $\geq 2$ hours/day | 94 (39.5) | 22 (28.2) | 11 (40.7) | 11 (36.7) | 0.336 |
| Sleep |  |  |  |  |  |
| Bedtime after 10 pm | 155 (65.1) | 44 (56.4) | 15 (55.6) | 25 (83.3) | **0.051** |
| Wake up after 7 am | 67 (28.2) | 20 (25.6) | 6 (22.2) | 9 (30.0) | 0.884 |
| Adequate sleep duration* | 160 (67.2) | 52 (66.7) | 18 (66.7) | 18 (60.0) | 0.890 |

Within parentheses are percentages over the column total for qualitative variables
Data were expressed in frequency (%) for qualitative and median (IQR) for quantitative variables
Chi-squared or Kruskal-Wallis test was done as appropriate

**Supplementary Table 6:** Physical and laboratory findings of the adult participants, n= 373.

| Variables | Control<br>n= 238 | Isolated irregular<br>cycle, n= 78 | Isolated<br>hirsutism<br>n= 27 | PCOS<br>n= 30 | p-value |
| --- | --- | --- | --- | --- | --- |
| BMI-status |  |  |  |  |  |
| Lean | 108 (45.4) | 38 (48.7) | 17 (63.0) | 16 (53.3) | 0.329 |
| Overweight-obese | 130 (54.6) | 40 (51.3) | 10 (37.0) | 14 (46.7) |  |
| Central obesity | 104 (43.7) | 36 (46.2) | 8 (29.6) | 16 (53.3) | 0.323 |
| MS-BP | 17 (7.1) | 8 (10.3) | 1 (3.7) | 2 (6.7) | 0.685 |
| Acanthosis nigricans | 32 (13.4) | 9 (11.5) | 3 (11.1) | 9 (30.0) |  |
| Goiter | 42 (17.6) | 19 (24.4) | 4 (14.8) | 0 (0.0)-<br>2.6 | **0.028** |
| Modified F-G score | 0.0 (0.0 –<br>1.0) | 1.0 (0.0 – 2.0) | 5.0 (4.0 –<br>9.0) | 4.0 (1.5 –<br>7.3) | **<0.001** |
| Acne | 36 (15.1) -<br>3.2 | 16 (20.5) | 13 (48.1) 3.8 | 10 (33.3) | **<0.001** |
| Glycemic status |  |  |  |  |  |
| NGT | 170 (71.4) | 46 (59.0) | 24 (88.9) | 19 (63.3) | **0.020** |
| AGT | 68 (28.6) | 32 (41.0) 2.3 | 3 (11.1) -2.3 | 11 (36.7) |  |
| MS-Triglyceride | 40 (16.8) | 19 (24.4) | 2 (7.4) | 9 (30.0) | 0.073 |
| MS-HDL cholesterol | 178 (74.8) | 63 (80.8) | 21 (77.8) | 25 (83.3) | 0.579 |
| Metabolic syndrome | 67 (28.2) | 24 (30.8) | 3 (11.1) | 10 (33.3) | 0.208 |
| MASH* | 17 (7.1) | 3 (3.8) | 2 (7.4) | 4 (13.3) |  |

Within parentheses are percentages over the column total for qualitative variables
Data were expressed in frequency (%) for qualitative and median (IQR) for quantitative variables
Chi-squared or Kruskal-Wallis test was done as appropriate

